# Delayed rise of oral fluid antibodies, elevated BMI, and absence of early fever correlate with longer time to SARS-CoV-2 RNA clearance in an longitudinally sampled cohort of COVID-19 outpatients

**DOI:** 10.1101/2021.03.02.21252420

**Authors:** Annukka A. R. Antar, Tong Yu, Nora Pisanic, Razvan Azamfirei, Jeffrey A. Tornheim, Diane M. Brown, Kate Kruczynski, Justin P. Hardick, Thelio Sewell, Minyoung Jang, Taylor Church, Samantha N. Walch, Carolyn Reuland, Vismaya S. Bachu, Kirsten Littlefield, Han-Sol Park, Rebecca L. Ursin, Abhinaya Ganesan, Oyinkansola Kusemiju, Brittany Barnaba, Curtisha Charles, Michelle Prizzi, Jaylynn R. Johnstone, Christine Payton, Weiwei Dai, Joelle Fuchs, Guido Massaccesi, Derek T. Armstrong, Jennifer L. Townsend, Sara C. Keller, Zoe O Demko, Chen Hu, Mei-Cheng Wang, Lauren M. Sauer, Heba H. Mostafa, Jeanne C. Keruly, Shruti H. Mehta, Sabra L. Klein, Andrea L. Cox, Andrew Pekosz, Christopher D. Heaney, David L. Thomas, Paul W. Blair, Yukari C. Manabe

**Affiliations:** Department of Medicine, Johns Hopkins University School of Medicine, Baltimore, MD, USA; Department of Environmental Health and Engineering, Johns Hopkins Bloomberg School of Public Health, Baltimore, MD, USA; W. Harry Feinstone Department of Molecular Microbiology and Immunology, Johns Hopkins Bloomberg School of Public Health, Baltimore, MD, USA; Department of Biochemistry and Molecular Biology, Johns Hopkins Bloomberg School of Public Health, Baltimore, MD, USA; Department of Pathology, Johns Hopkins University School of Medicine, Baltimore, MD, USA; Department of Oncology, Johns Hopkins University School of Medicine, Baltimore, MD, USA; Department of Biostatistics, Johns Hopkins Bloomberg School of Public Health, Baltimore, MD, USA; Department of Emergency Medicine, Johns Hopkins University School of Medicine, Baltimore, MD, USA; Department of Epidemiology, Johns Hopkins Bloomberg School of Public Health, Baltimore, MD, USA; Department of International Health, Johns Hopkins Bloomberg School of Public Health, Baltimore, MD, USA

## Abstract

**Background:** Sustained molecular detection of SARS-CoV-2 RNA in the upper respiratory tract (URT) in mild to moderate COVID-19 is common. We sought to identify host and immune determinants of prolonged SARS-CoV-2 RNA detection.

**Methods:** Ninety-five outpatients self-collected mid-turbinate nasal, oropharyngeal (OP), and gingival crevicular fluid (oral fluid) samples at home and in a research clinic a median of 6 times over 1-3 months. Samples were tested for viral RNA, virus culture, and SARS-CoV-2 and other human coronavirus antibodies, and associations were estimated using Cox proportional hazards models.

**Results:** Viral RNA clearance, as measured by SARS-CoV-2 RT-PCR, in 507 URT samples occurred a median (IQR) 33.5 (17-63.5) days post-symptom onset. Sixteen nasal-OP samples collected 2-11 days post-symptom onset were virus culture positive out of 183 RT-PCR positive samples tested. All participants but one with positive virus culture were negative for concomitant oral fluid anti-SARS-CoV-2 antibodies. The mean time to first antibody detection in oral fluid was 8-13 days post-symptom onset. A longer time to first detection of oral fluid anti-SARS-CoV-2 S antibodies (aHR 0.96, 95% CI 0.92-0.99, p=0.020) and BMI ≥ 25kg/m^2^ (aHR 0.37, 95% CI 0.18-0.78, p=0.009) were independently associated with a longer time to SARS-CoV-2 viral RNA clearance. Fever as one of first three COVID-19 symptoms correlated with shorter time to viral RNA clearance (aHR 2.06, 95% CI 1.02-4.18, p=0.044).

**Conclusions:** We demonstrate that delayed rise of oral fluid SARS-CoV-2-specific antibodies, elevated BMI, and absence of early fever are independently associated with delayed URT viral RNA clearance.

## INTRODUCTION

Although severe acute respiratory syndrome coronavirus 2 (SARS-CoV-2) infections chiefly occur in the community, early virus and host response kinetics have been largely described in hospitalized inpatients. The logistics of recruiting and sampling potentially infectious individuals in the outpatient setting have sustained gaps in our knowledge of infectivity and pathogenesis in mild to moderate disease. In particular, it is not known whether antibody kinetics are tightly correlated on an individual level with viral RNA decay in mild to moderate COVID-19. Several case reports of prolonged shedding of infectious virus in immune compromised people with COVID-19[1-4] support the hypothesis that adaptive immunity is required for clearance of infectious virus. Two studies report that prolonged viral RNA shedding is correlated with lower peak SARS-CoV-2-specific IgG titers and lower numbers of B and T cells in blood[5, 6]. However, other studies report no difference in plasma SARS-CoV-2-specific antibody titers in patients testing positive or negative at late timepoints after symptom onset[7, 8]. Here we leveraged guided home sampling to longitudinally characterize viral and immune kinetics in a prospective cohort with mild to moderate COVID-19 and identified associations with duration of viral RNA shedding from the upper respiratory tract that yield insights into COVID-19 pathogenesis.

## METHODS

### Study Cohort and Sampling

The Johns Hopkins University (JHU) School of Medicine Institutional Review Board approved this study. Participants provided informed consent. The study prospectively enrolled a convenience sample of adults ≥ 30 years old within 48 hours of a positive SARS-CoV-2 reverse-transcriptase polymerase chain reaction (RT-PCR) test from an outpatient testing site of the Johns Hopkins Health System (JHHS) between April 21, 2020, and July 23, 2020 (Figure 1A)[9, 10]. The 95 participants with known symptom onset date and who completed one or more RT-PCR tests after symptom onset were included here. Self-sampled mid-turbinate nasal swabs and oropharyngeal (OP) swabs were obtained with telephone guidance from study staff, and swabs were combined in 3mL viral transport medium and frozen. Mid-turbinate nasal-OP RT-PCR is equivalent to clinician-collected nasopharyngeal swabs[11-15]. Gingival crevicular fluid was obtained using the Oracol device (Malvern Medical Developments Ltd., Worcestershire, UK). Saliva was collected in the tube from June 1, 2020, onwards after validation of the spit saliva sample type[1]. The sampling schedule is shown in Figure 1B. Fifty-three of 95 participants presented to a research clinic visit at 1-4 months post-symptom onset for height, weight, vital signs, blood draw, nasopharyngeal swab, and oral fluid sampling. Symptom reports were collected on the same days as sample collection.

**Figure 1.**
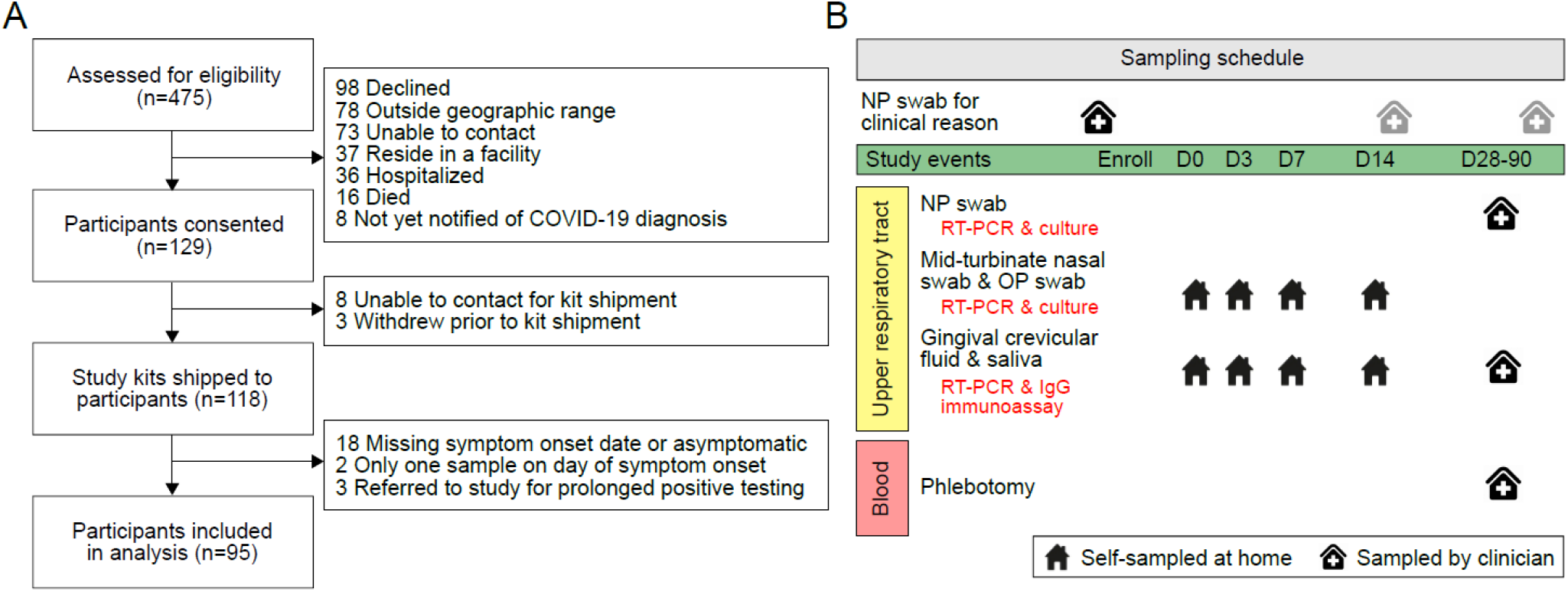
CONSORT flow diagram and sampling schedule. (A) A convenience sample of 475 adults with recent positive SARS-CoV-2 RT-PCR test from an outpatient testing site of the Johns Hopkins Health System was assessed for eligibility between April 21, 2020, and July 23, 2020. Preference was given to participants 40 years of age and older. Study kits were sent to 118 participants, and data and samples from 95 participants were included in the analyses presented here. (B) Study sampling schedule. Clinical RT-PCR results from the medical record are included in these analyses. Samples on study days 0-14 were self-collected at home with telephone or video guidance by trained study staff. Participants presented to a research clinic for collection of samples a median of 45 days after study day 0 (range 27-88).

### Laboratory Procedures

RT-PCR testing of study samples was performed on the Abbott *m*2000 platform (Abbott Molecular, Des Plaines, Il) in 600 µl volumes[9]. Reported are cycle threshold (Ct) values from one of two primer/probe regions. Ct values < 31.5 was positive. 200 µl volumes of oral fluid were assayed by RT-PCR. The adequacy of self-collected nasal-OP and oral fluid samples was confirmed via quantitative PCR for GAPDH human gene expression utilizing a TaqMan gene expression assay (ThermoFisher Scientific, Waltham, MA) (data not shown).

Positive nasal-OP samples by RT-PCR were tested for propagation of SARS-CoV-2 in cell culture[16]. Plasma SARS-CoV-2-specific antibody titers were quantified by indirect ELISA and neutralizing antibody titers were quantified by microneutralization assay[17, 18]. Quantification of oral fluid SARS-CoV-2-specific IgG and other human coronavirus-specific IgG was obtained from a multiplex SARS-CoV-2 immunoassay based on Luminex technology[19, 20]. The oral fluid SARS-CoV-2-specific IgG readout of this assay is highly correlated with plasma SARS-CoV-2 neutralizing antibodies and plasma S-RBD-specific IgG by ELISA[19, 20].

### Statistical methods

Kaplan-Meier plots and log-rank were generated using the ‘survival’ R package[21]. Associations between covariates and clearance of upper respiratory tract SARS-CoV-2 viral RNA was determined via Cox proportional hazards models[21] after multiple imputation and model selection by least absolute shrinkage and selection operator (LASSO). A complete list of covariates for Cox model 1 may be found in Supplementary Table 2. Time to RT-PCR clearance was defined as number of days from symptom onset to the midpoint between the last positive RT-PCR test and the subsequent negative test. Twenty datasets were imputed using predictive mean matching for univariate imputation and chained equation (MICE) for multivariate imputation using the ‘mice’ R package[22]. Model selection was performed using LASSO on each of the imputed datasets using the ‘glmnet’ R package[23]. Nine variables were included in the model, which used the pooled imputed dataset (Figure 5, Supplementary Table 2). To determine whether the time from symptom onset to first detection of oral fluid SARS-CoV-2-specific IgG is associated with RT-PCR clearance, we performed model selection using LASSO and ran Cox regression model 2. Further details provided in Supplementary Methods.

## RESULTS

### Study design

Ninety-five non-hospitalized individuals with a positive nasopharyngeal SARS-CoV-2 RT-PCR test from an emergency room or an ambulatory testing center within the previous 48 hours were enrolled between April 21, 2020, and July 23, 2020 (Figure 1A). Study day 0 was a median of 4 days (range 2-11) from the collection of the most recent prior positive RT-PCR test and was a median of 9 days from symptom onset (range 2-80). Participants presented for a single research clinic visit a median of 45 days after study day 0 (range 27-88).

### Clinical characteristics

The median age of the participants was 56 years, and 59% were women (Table 1). A total of 39% identified as Black or African American, and 14% as Hispanic/LatinX. The median body mass index (BMI) of participants was 29.3 kg/m^2^, similar to that of US adults, 29.4 kg/m^2^ [24]. Eight participants (8.4%) required hospitalization after enrollment in the study[10].

**Table 1.**
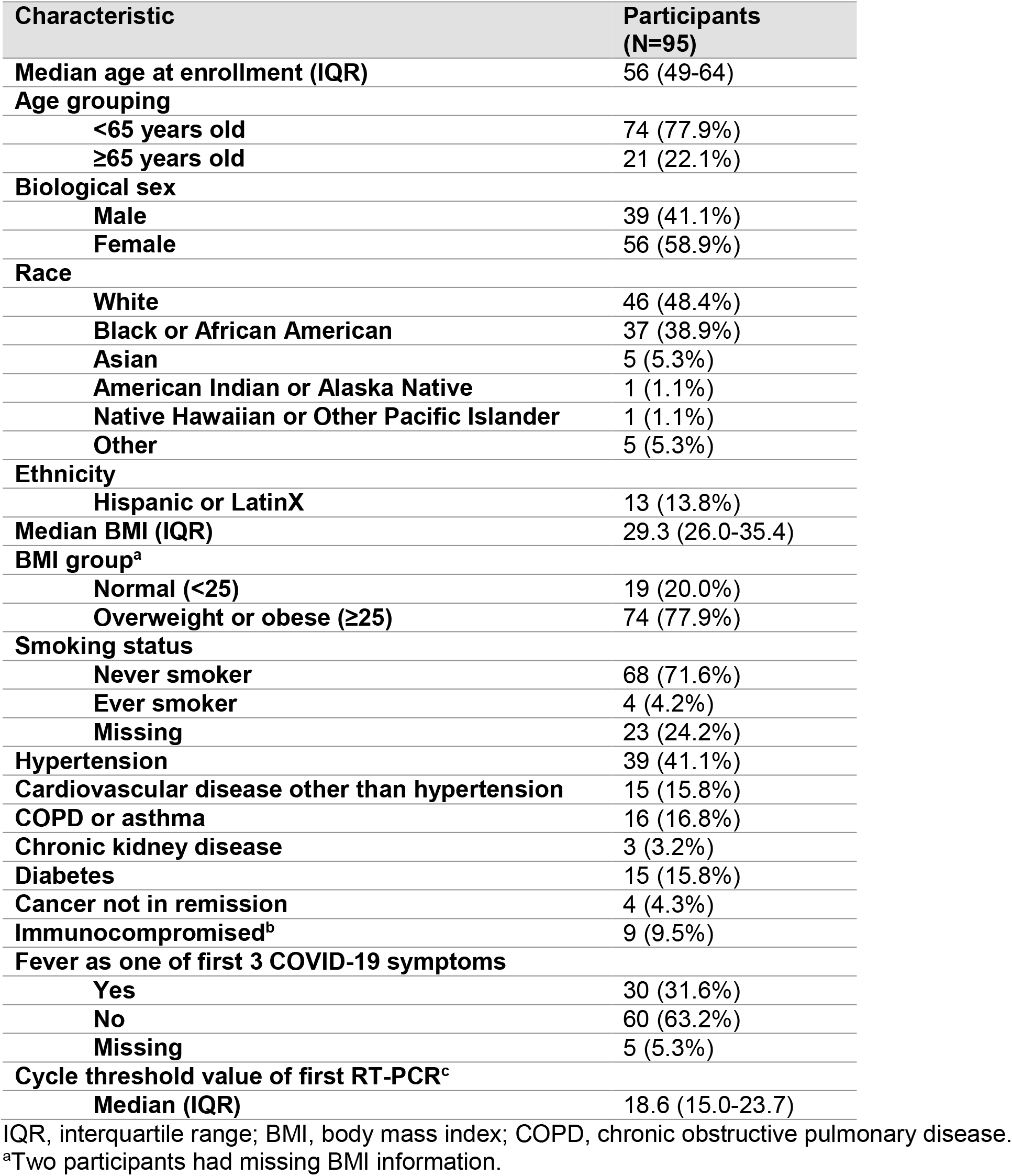
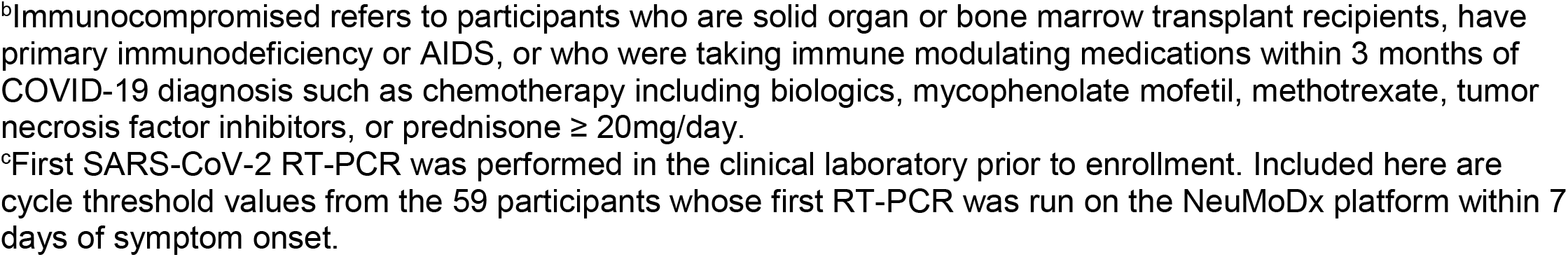
Demographics and clinical characteristics of participants.

| Characteristic | Participants (N=95) |
| --- | --- |
| **Median age at enrollment (IQR)** | 56 (49-64) |
| **Age grouping** |  |
| <65 years old | 74 (77.9%) |
| ≥65 years old | 21 (22.1%) |
| **Biological sex** |  |
| Male | 39 (41.1%) |
| Female | 56 (58.9%) |
| **Race** |  |
| White | 46 (48.4%) |
| Black or African American | 37 (38.9%) |
| Asian | 5 (5.3%) |
| American Indian or Alaska Native | 1 (1.1%) |
| Native Hawaiian or Other Pacific Islander | 1 (1.1%) |
| Other | 5 (5.3%) |
| **Ethnicity** |  |
| Hispanic or LatinX | 13 (13.8%) |
| **Median BMI (IQR)** | 29.3 (26.0-35.4) |
| **BMI group<sup>a</sup>** |  |
| Normal (<25) | 19 (20.0%) |
| Overweight or obese (≥25) | 74 (77.9%) |
| **Smoking status** |  |
| Never smoker | 68 (71.6%) |
| Ever smoker | 4 (4.2%) |
| Missing | 23 (24.2%) |
| **Hypertension** | 39 (41.1%) |
| **Cardiovascular disease other than hypertension** | 15 (15.8%) |
| **COPD or asthma** | 16 (16.8%) |
| **Chronic kidney disease** | 3 (3.2%) |
| **Diabetes** | 15 (15.8%) |
| **Cancer not in remission** | 4 (4.3%) |
| **Immunocompromised<sup>b</sup>** | 9 (9.5%) |
| **Fever as one of first 3 COVID-19 symptoms** |  |
| Yes | 30 (31.6%) |
| No | 60 (63.2%) |
| Missing | 5 (5.3%) |
| **Cycle threshold value of first RT-PCR<sup>c</sup>** |  |
| Median (IQR) | 18.6 (15.0-23.7) |

IQR, interquartile range; BMI, body mass index; COPD, chronic obstructive pulmonary disease.
<sup>a</sup>Two participants had missing BMI information.
<sup>b</sup>Immunocompromised refers to participants who are solid organ or bone marrow transplant recipients, have primary immunodeficiency or AIDS, or who were taking immune modulating medications within 3 months of COVID-19 diagnosis such as chemotherapy including biologics, mycophenolate mofetil, methotrexate, tumor necrosis factor inhibitors, or prednisone $\geq 20\text{mg/day}$ .
<sup>c</sup>First SARS-CoV-2 RT-PCR was performed in the clinical laboratory prior to enrollment. Included here are cycle threshold values from the 59 participants whose first RT-PCR was run on the NeuMoDx platform within 7 days of symptom onset.

**Table 2.** Time-dependent covariate Cox proportional hazards model for viral RNA clearance (Cox model 2).

| Variable | HR | aHR | 95% CI | p value |
| --- | --- | --- | --- | --- |
| Change from undetectable to detectable Mt. Sinai S-specific oral fluid IgG | 0.96 | 0.96 | 0.92 – 0.99 | **0.020** |
| BMI $\geq 25 \text{ kg/m}^2$ (referent: BMI < 25) | 0.44 | 0.29 | 0.14 – 0.61 | **0.001** |
| Diabetes | 2.11 | 2.49 | 1.09 – 5.67 | **0.030** |
| Immunocompromised | 0.68 | 0.35 | 0.12 – 1.02 | 0.055 |
| Fever as one of first 3 COVID-19 symptoms | 1.72 | 2.23 | 1.05 – 4.71 | **0.036** |
| **Race/ethnicity (referent: non-Hispanic White)** |  |  |  |  |
| African American or Black race, any ethnicity | 0.73 | 0.85 | 0.42 – 1.73 | 0.659 |
| Non-White, Non-Black, non-Hispanic | 1.20 | 0.56 | 0.14 – 2.23 | 0.408 |
| Hispanic of any race except Black | 0.79 | 1.48 | 0.41 – 5.39 | 0.549 |

HR, hazard ratio; aHR, adjusted hazard ratio; CI, confidence interval; S, SARS-CoV-2 spike protein

### Viral kinetics in COVID-19 outpatients

Participants’ SARS-CoV-2 RT-PCR tests from the study and from the medical record were included in these analyses, totaling 507 RT-PCR tests (Figure 2). Mid-turbinate nasal-OP and nasopharyngeal RT-PCR tests are the primary sample types, while oral fluid RT-PCR tests were included only if nasal samples were missing or in the 3 instances in which the nasal-OP sample was negative while the oral fluid sample was positive. Through December 15, 2020, the 95 participants had a median of 6 (IQR 4-6) RT-PCR tests during and after acute COVID-19. Longitudinal Ct values from study samples showed a rapid decay with several instances of a negative test followed by a positive test (re-positive cases) (Figure 3A, gray lines). To examine Ct values as an indicator of viral load in the URT from very early timepoints, RT-PCR Ct values were obtained from the participant’s first positive clinical RT-PCR test, the majority of which were run on the NeuMoDx platform. The mean Ct value was lowest - meaning the viral burden was highest - before and just after symptom onset (Figure 3B). The first 8-10 cycles on the Abbott RT-PCR platform are not read, and so the Ct values of the Abbott (study samples) and NeuMoDx (clinical samples) platforms are not comparable.

**Figure 2.**
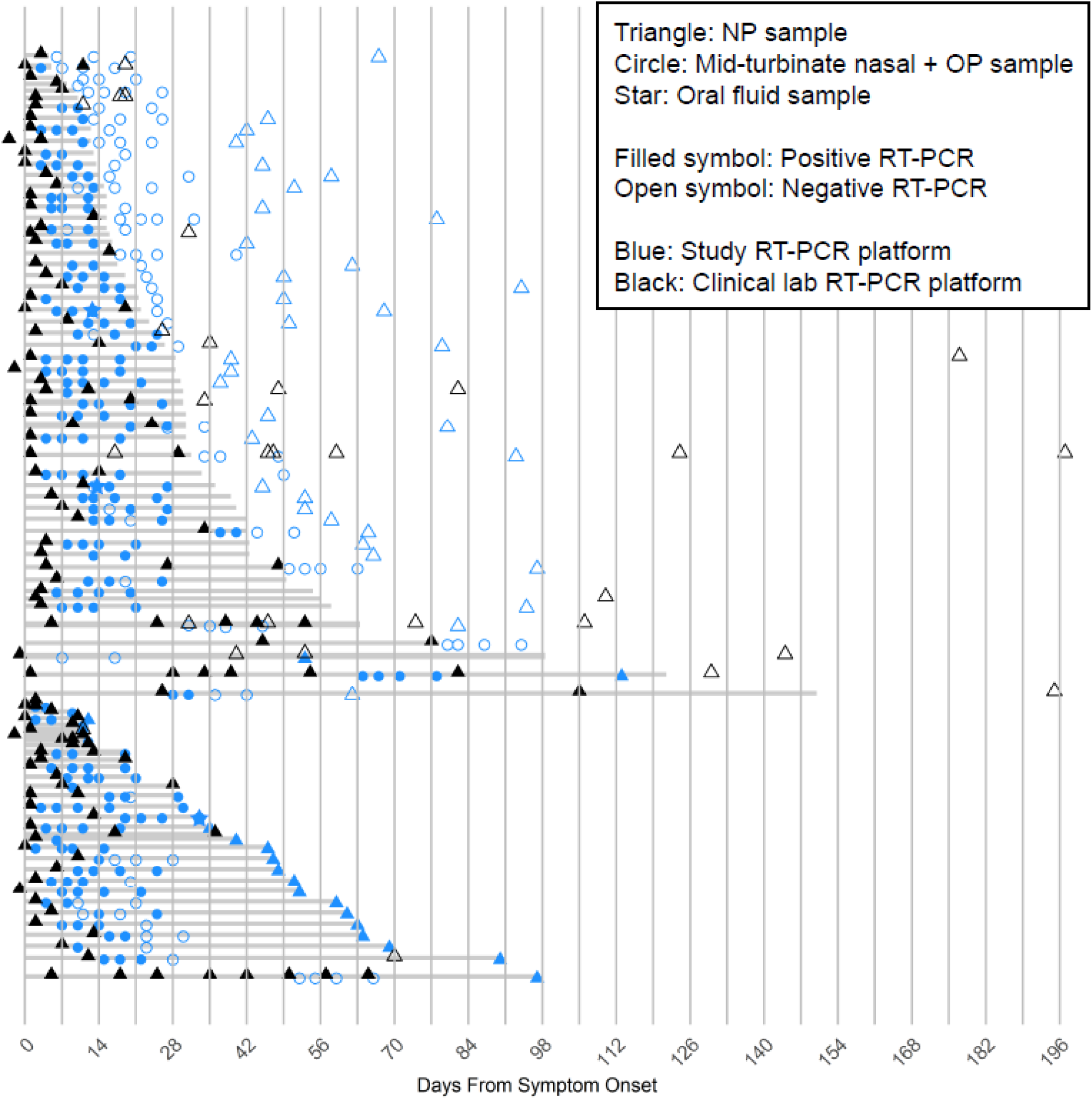
Longitudinal SARS-CoV-2 RT-PCR sampling in outpatients. SARS-CoV-2 RT-PCR test results by day from symptom onset. Each row represents one participant. Shown here are nasal samples. Stars indicate a sample timepoint at which the nasal sample was negative and the oral fluid sample was positive. Included in the top grouping are participants whose last RT-PCR test was negative, and in the bottom group, those whose last RT-PCR test was positive. Shaded gray lines indicate the number of days from symptom onset until the mid-point between the last positive sample and the next negative sample (top grouping) or until the last positive sample (bottom grouping). Nasopharyngeal (NP) swabs collected by a health care worker are indicated by triangles, self-collected mid-turbinate nasal-oropharyngeal (OP) swab combined in viral transport media are indicated by circles.

**Figure 3.**
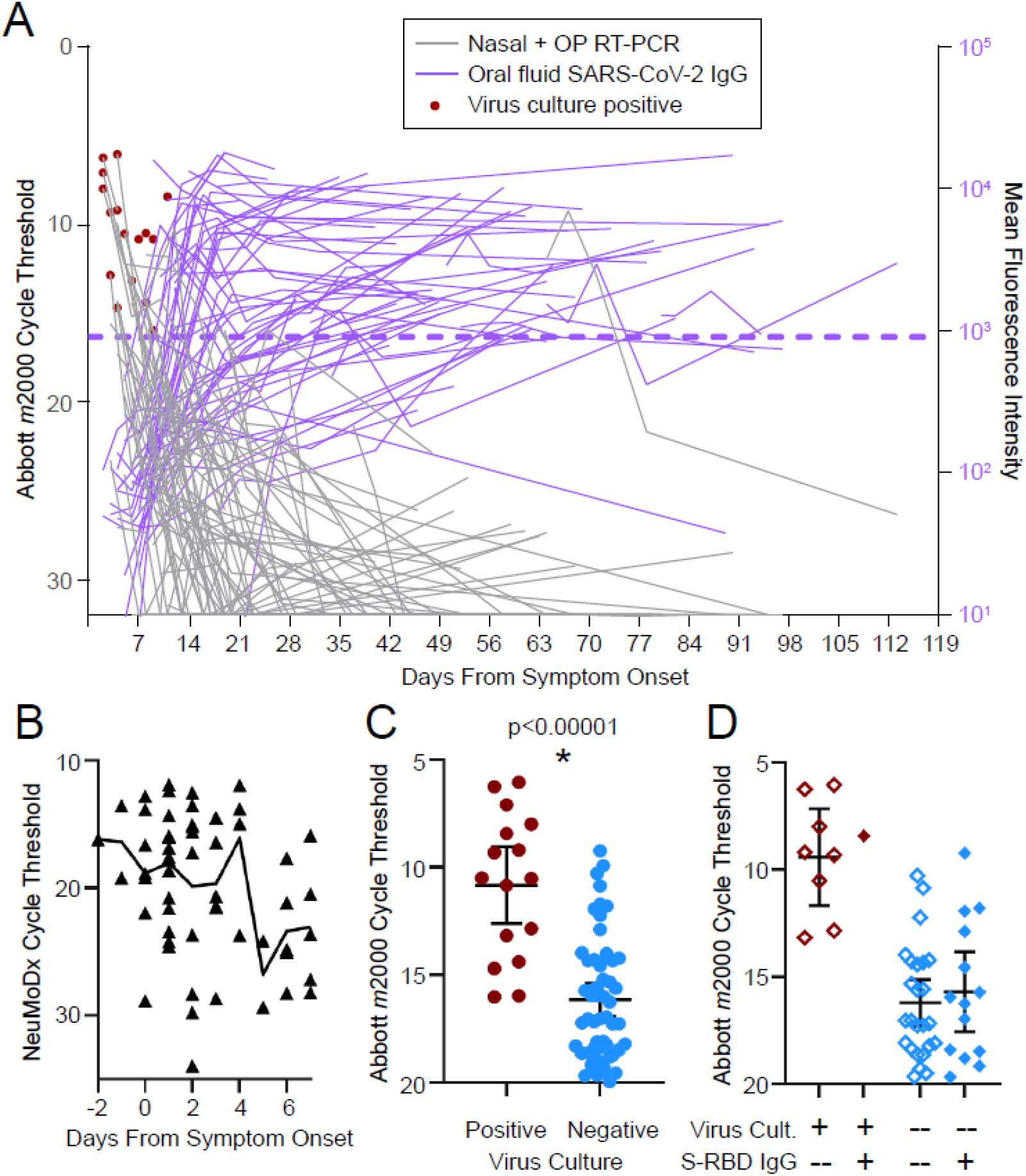
Kinetics of RT-PCR cycle thresholds, virus culture, and oral fluid SARS-CoV-2 antibodies in mild-moderate COVID-19. (A) Longitudinal Abbott *m*2000 SARS-CoV-2 RT-PCR cycle threshold values from nasal-oropharyngeal (OP) swab study samples (blue circles and triangles in Fig. 2) by day from symptom onset are shown in gray. Longitudinal oral fluid anti-spike (Mt. Sinai) IgG mean fluorescence intensity shown in purple. Lines connect values from the same participant. Each sample with a positive RT-PCR result was assessed for propagation of SARS-CoV-2 on VeroE6-TMPRSS2 cells. Samples with positive virus culture are indicated by red circles. The cutoff for positive anti-S IgG is shown with a purple dotted line (see Methods for cutoff calculation). (B) The subset of 59 participants with a nasopharyngeal swab collected within 7 days of symptom onset and run on the NeuMoDx SARS-CoV-2 RT-PCR platform are shown by day from symptom onset. Line connects cycle threshold means. (C) The subset of nasal-OP swab samples with cycle threshold < 20 is grouped according to whether virus culture of the sample was positive or negative. Means with 95% confidence intervals shown. * indicates p < 0.00001 by one-tailed student’s t test. (D) The subset of the samples shown in (B) that had a simultaneous adequate oral fluid sample are shown here. Filled diamonds represent study timepoints at which oral fluid contained detectable IgG antibodies to Mt Sinai S-RBD, and open diamonds represent study timepoints at which oral fluid was negative for anti-S-RBD IgG. Cult, culture; S-RBD, spike receptor binding domain; Ab, antibody.

### Virus culture positivity was detected through 11 days post-symptom onset

Sixteen (of 183 RT-PCR positive) study samples cultured for SARS-CoV-2 on VeroTMPRSS2 cells[16, 25] from 14 participants were virus culture positive (Figure 3A, red circles). In these 14 participants, the median time from symptom onset to last positive virus culture was 5 days (range 2-11). No sample tested positive for virus culture beyond 11 days from symptom onset, even in the case of an immunocompromised participant who had positive RT-PCRs with low Ct values two months post-symptom onset (Figure 3A). None of the re-positive RT-PCR samples were positive for virus culture. Virus culture was positive only in samples with Ct values <17 on the Abbott *m*2000 platform. The mean Ct value of samples positive for virus culture was significantly lower than that of samples negative for viral culture (Figure 3C).

### Positive oral fluid SARS-CoV-2-specific IgG is associated with negative virus culture

To determine whether oral fluid anti-SARS-CoV-2 IgG could be used to predict which samples with low Ct values were negative for virus culture, we examined sample timepoints with (1) nasal Abbott RT-PCR Ct values < 20, (2) nasal virus culture results, and (3) oral fluid samples with adequate total IgG for assessment (total IgG >10 µg/mL or detectable oral fluid anti-S-RBD IgG). Fourteen of 15 samples positive for oral fluid anti-S-RBD IgG were negative for virus culture (Figure 3D). The one culture positive sample was collected on day 11 after symptom onset, which is at or shortly after the expected first detection of this antibody (Supplementary Table 1).

### Oral fluid SARS-CoV-2-specific IgG is first detected between 8 and 13 days from symptom onset

Oral fluid SARS-CoV-2-specific IgG to multiple viral antigens was assessed (Supplementary Figures 1-3). Longitudinal kinetics of oral fluid anti-SARS-CoV-2 S IgG by day from symptom onset are shown in purple in Figure 3A. Supplementary Table 1 shows the means and 95% confidence intervals of number of days from symptom onset to rise above cutoff for each of the eight SARS-CoV-2 antigens (8-13 days depending on the antigen), similar to that previously described[19].

### Kinetics of SARS-CoV-2 viral RNA clearance in outpatients

The median time to clearance was 33.5 days (range 4.5-150, IQR 17-63.5; Figure 4A), similar to hospitalized COVID-19 patients with a similar mean age[26].

**Figure 4.**
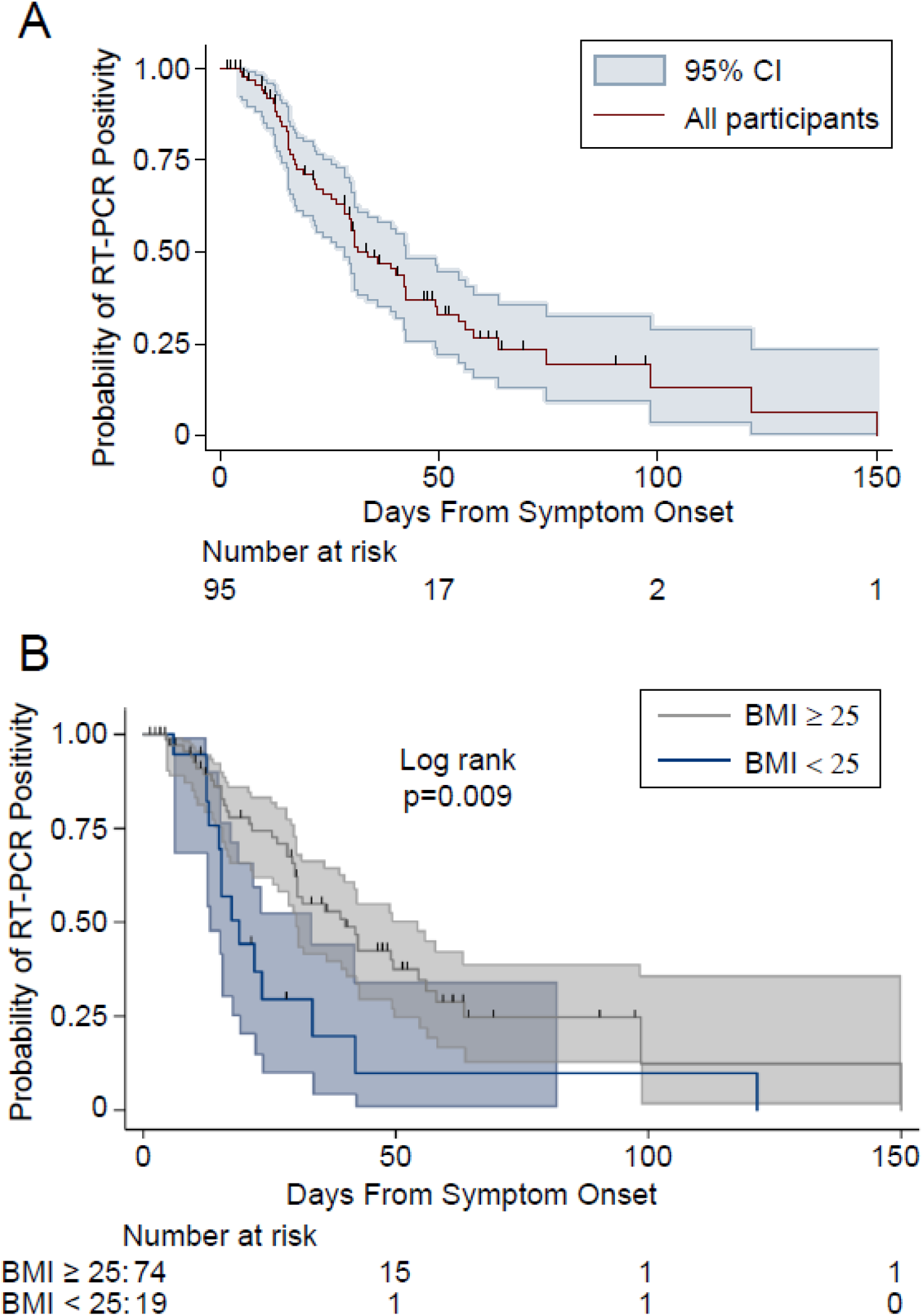
Kaplan Meier plots SARS-CoV-2 RT-PCR positivity by day from symptom onset in mild-moderate COVID-19. (A) Kaplan Meier survival curve with 95% confidence intervals (CI) for positive upper respiratory tract SARS-CoV-2 RT-PCR by day from symptom onset. (B) Kaplan Meier survival curve by BMI category.

### Elevated BMI is associated with longer time to viral RNA clearance

To determine host and immune factors associated with longer time to RT-PCR clearance, we used a Cox proportional hazards model after multiple imputation and model selection by LASSO. Nine covariates were selected for inclusion in the Cox model. Shown in Figure 5 are the univariable and adjusted hazard ratios of clearance and the 95% confidence intervals. BMI ≥ 25kg/m^2^ was independently associated with longer time to viral RNA clearance in outpatients with COVID-19 (aHR 0.37, 95% CI 0.18-0.78, p=0.009) (Figures 4B & 5). This association did not appear to be explained by comorbidities such as hypertension and diabetes as inclusion of hypertension and diabetes in the model did not attenuate the BMI association. Further, hypertension was not significantly associated with time to viral RNA clearance and diabetes was significantly positively associated with clearance, which is in the opposite direction of elevated BMI.

**Figure 5.**
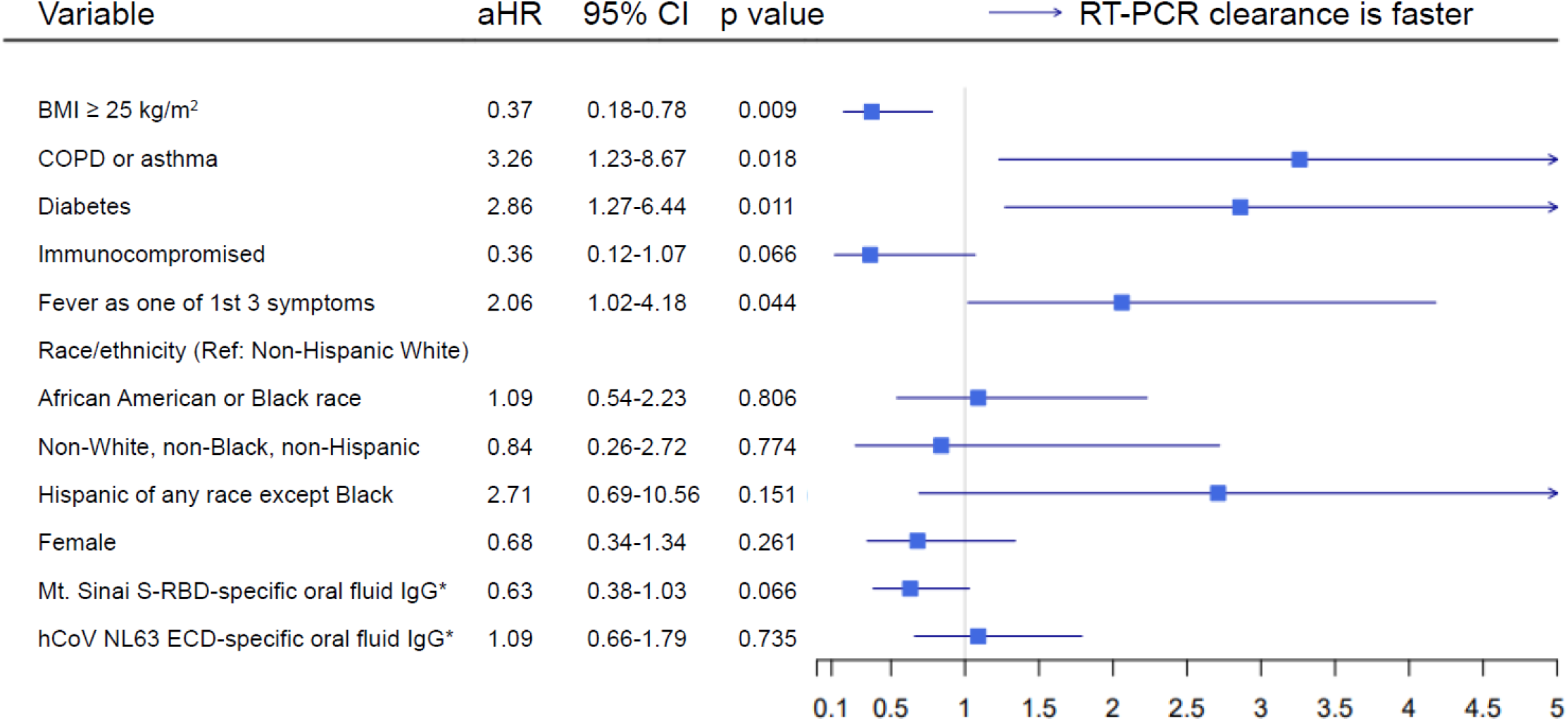
Cox proportional hazards model for viral RNA clearance. An adjusted hazard ratio (aHR) < 1 denotes longer time to SARS-CoV-2 viral RNA clearance in the upper respiratory tract as measured by RT-PCR. To account for missing datapoints, 20 datasets were imputed using predictive mean matching (PPM) multiple imputation by chained equation (MICE). Model selection using least absolute shrinkage and selection operator (LASSO) was performed on each of the 20 datasets. The top 9 variables of 35 - selected 15 or more times out of 20 (Supplementary Table 2) - were chosen for inclusion in the Cox proportional hazards model, which was performed using the pooled imputed dataset. *, standardized log mean fluorescence intensity and treated as a time-varying variable. S-RBD, spike receptor binding domain; hCoV, human coronavirus; ECD, spike ectodomain.

There was a trend towards immunocompromised hosts (n=9) having a longer time to viral RNA clearance (Figure 5). There was additionally a trend towards greater magnitude of oral fluid anti-S-RBD IgG being associated with longer time to RT-PCR clearance (Figure 5). The magnitude of oral fluid anti-human coronavirus IgG for the four circulating non-SARS human coronaviruses was not associated with time to viral RNA clearance (Figure 5, Supplementary Table 2).

### COPD/asthma, early fever, and diabetes are associated with faster viral RNA clearance

COPD/asthma, diabetes, and fever reported as one of first three COVID-19 symptoms were each associated with faster viral RNA clearance (Figure 5). In a sensitivity analysis excluding participants who cleared ≥ 75 days after symptom onset, only BMI and COPD/asthma were still significantly associated (Supplementary Table 5).

### Longer time to first detection of oral fluid SARS-CoV-2-specific IgG is associated with longer time to viral RNA clearance

In a time-dependent covariate Cox model (Cox model 2) for viral RNA clearance, a longer time to first detection of oral fluid spike-specific IgG is associated with longer time to viral RNA clearance (aHR 0.96, 95% CI 0.92-0.99, p=0.020).

### Plasma SARS-CoV-2-specific antibody titer in convalescence is not associated with time to viral RNA clearance

Titers of plasma anti-S-RBD IgG, anti-S IgG, or neutralizing antibody in convalescence was not significantly associated with viral RNA clearance in either direction. Additionally, the presence or absence of viral RNA by RT-PCR in the nasopharynx at 1-4 months post-symptom onset was not associated with plasma antibody titer (data not shown).

## DISCUSSION

In a racially- and ethnically-diverse cohort of 95 adult outpatients, we report that a longer time to first detection of oral fluid SARS-CoV2-specific antibodies is independently associated with a longer time to viral RNA clearance. We find that the presence of SARS-CoV-2 antibodies in oral fluid may be a predictor of negative virus culture from nasal samples, even in samples with low RT-PCR Ct values. We demonstrate that the mean time from symptom onset to first detection of oral fluid SARS-CoV-2 S-RBD-IgG is 9-11 days, and virus culture positive samples may be detected through day 11 post-symptom onset in adults with mild-to-moderate COVID-19, one day longer than the current recommended isolation period[27].

The strengths of this cohort are its size, its prospective and intensive longitudinal sampling design beginning in the acute phase of illness, its long duration of follow-up, and its simultaneous testing of samples for SARS-CoV-2 RT-PCR, virus culture, and oral fluid antibodies. Two studies of mostly outpatients included more cultured specimens but were not longitudinal[28, 29]. Our findings extend to the outpatient domain those described in 129 patients with severe COVID-19 by van Kampen and colleagues[30], who report mean time from symptom onset to last positive virus culture of 8 days (maximum 20), and that virus cultures were more likely to be negative when plasma neutralizing antibodies were elevated[30]. Our results suggest that outpatients testing positive for SARS-CoV-2 who have detectable oral fluid SARS-CoV-2-specfic antibodies are not likely to be transmissible, but larger studies are needed for confirmation.

We demonstrate that elevated BMI is also independently associated with longer time to viral RNA clearance. This association is not likely explained by increased prevalence of comorbidities such as hypertension or diabetes. In fact, diabetes, along with early fever and COPD/asthma, is associated with faster time to viral RNA clearance. Numerous studies report strong associations of COVID-19 severity and older age with viral RNA shedding duration[17, 31, 32]. Neither severity nor age were significantly associated with duration of viral RNA shedding in our study, likely because the vast majority of our cohort participants were in the same severity category and age range (IQR 49-64). Importantly, we found no effect of race or ethnicity with time to viral RNA clearance after adjustment for other demographic and comorbidities. Obesity is associated with testing positive for SARS-CoV-2, with severity of COVID-19, and with COVID-19 mortality[33-36]. Dysregulated immunity is observed in severe COVID-19, aging, and obesity, and it may be the unmeasured key correlate of viral RNA shedding duration. Aging drastically increases the chronic state of inflammation present in obesity[37], perhaps explaining why our study of middle-aged and older participants uncovered the strong association of elevated BMI and viral RNA shedding duration. The other risk factors that correlated with longer time to viral RNA clearance in our study – absence of early fever and longer time to detection of oral fluid antibodies – are consistent with impaired early innate and adaptive immune responses. Silva et al. report that a delay in antibody production is linked to high salivary viral loads in hospitalized patients[38]. We hypothesize that the chronic inflammatory state of aging and obesity hinders early innate and adaptive immune responses to SARS-CoV-2, leading to higher viral loads which require higher antibody titers and more time to clear, while also presenting a cytokine milieu in which it is stochastically more likely for a patient to tip into the positive feedback loops of hyperinflammation that portend disease severity. Mechanistic studies are needed to determine how obesity prolongs viral RNA shedding.

Our study has limitations, including that it is under-sampled at the time of symptom onset and at > 1 month from symptom onset. We address this by including clinician-ordered samples outside of the study, but the time to viral RNA clearance may be less precisely described in participants whose study samples at 1-3 months remained positive than others. There was a non-significant trend towards immunocompromised host status being associated with viral RNA shedding duration, which may appear to contrast with reports of prolonged viral carriage in immunocompromised people[1-4]. This likely occurred because we grouped all immunocompromised people together, whereas there is probably a specific immunocompromised phenotype that leads to prolonged viral RNA detection. The associations of diabetes and COPD/asthma with faster time to viral RNA clearance were not found to be independent of other covariates in our sensitivity analysis (diabetes) or our second Cox model (COPD/asthma), and conflict with one report of asthma being associated with longer viral RNA shedding[39]. Larger studies are needed to determine whether diabetes, COPD, or asthma are associated with time to viral RNA clearance.

In sum, we report viral and immune kinetics in an intensively characterized cohort of 95 adult outpatients with mild to moderate COVID-19. We demonstrate that elevated BMI, longer time to detection of oral SARS-CoV-2 IgG, and the absence of early fever are independently associated with longer time to viral RNA clearance, suggesting that functional early innate and adaptive immunity is critical for timely viral RNA clearance.

## Data Availability

De-identified data is not currently available, but can be made available upon request.

## Acknowledgements

We acknowledge Jeffrey Holden, Tyrone Howard, Andrew Karaba, Anastasia Lambrou, Lucy Li, Manuela Plazas Montana, Caroline Popper, and Amanda Tuchler for their assistance.

