## Supplementary Data and Methods for "Delayed rise of oral fluid antibodies, elevated BMI, and absence of early fever correlate with longer time to SARS-CoV-2 RNA clearance in an longitudinally sampled cohort of COVID-19 outpatients"

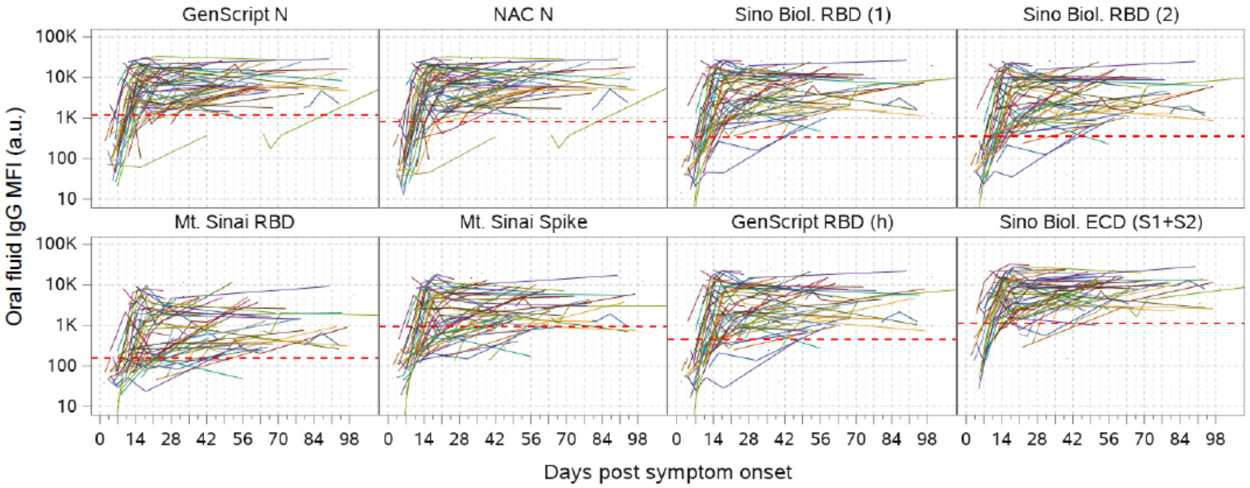


**Supplementary Figure 1. Oral fluid SARS-CoV-2-specific IgG kinetics in mild to moderate COVID-19.** Gingival crevicular fluid (oral fluid) SARS-CoV-2-specific IgG levels by day from symptom onset in participants detected using an in-house multiplex SARS-CoV-2 immunoassay based on Luminex technology as described previously[1,2]. Lines connect values from the same participant. Cutoffs for each SARS-CoV-2 antigen were defined as the average net (blank-subtracted) MFI of pre-COVID-19 era oral fluid samples plus three standard deviations (SD) and are denoted by red dotted lines. To minimize false negatives due to low concentration of total IgG, samples with less than 15 μg/mL total oral fluid IgG are not shown here unless the sum of the signal to cutoff ratio for 7 SARS-CoV-2 antigens was above 6. MFI, mean fluorescence intensity; a.u., arbitrary units; N, nucleocapsid; NAC, Native Antigen Company; RBD, receptor binding domain of spike protein; ECD, spike ectodomain; S1, spike subunit 1; S2, spike subunit 2.

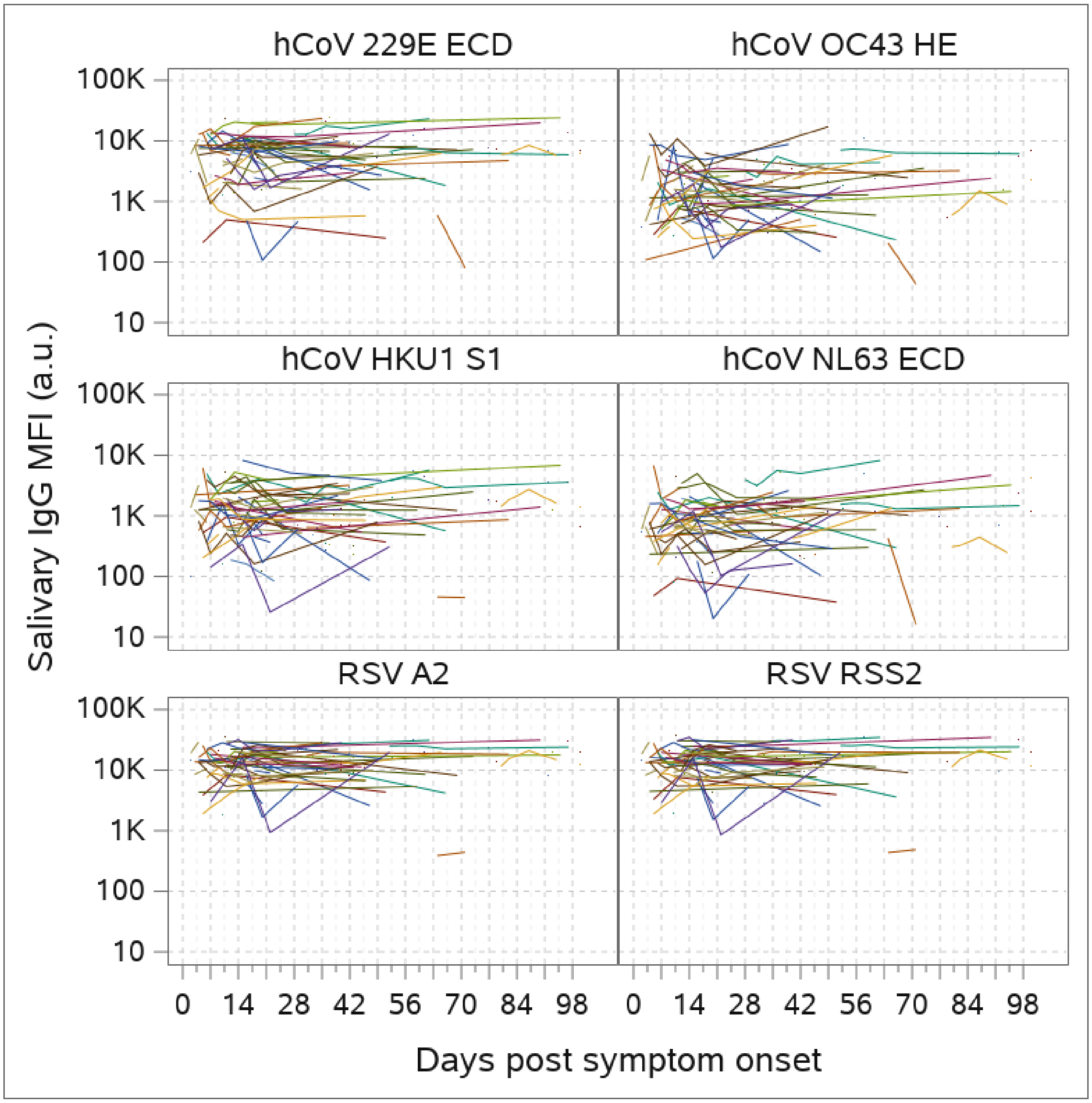


**Supplementary Figure 2. Non-SARS-CoV-2 virus-specific IgG kinetics in oral fluid.** Results shown are from an in-house multiplex SARS-CoV-2 immunoassay based on Luminex technology. Lines connect values from the same participant. To minimize false negatives due to low concentration of total antibody, samples with less than 15 µg/mL total oral IgG are not shown here unless they were also positive for most SARS-CoV-2 specific antigens. MFI, mean fluorescence intensity; a.u. arbitrary units; hCoV, human coronavirus; ECD, spike ectodomain; HE, hemagglutinin esterase; S1, spike subunit 1; RSV, respiratory syncytial virus; A2, A2 strain of RSV; RSS2, RSS2 strain of RSV.

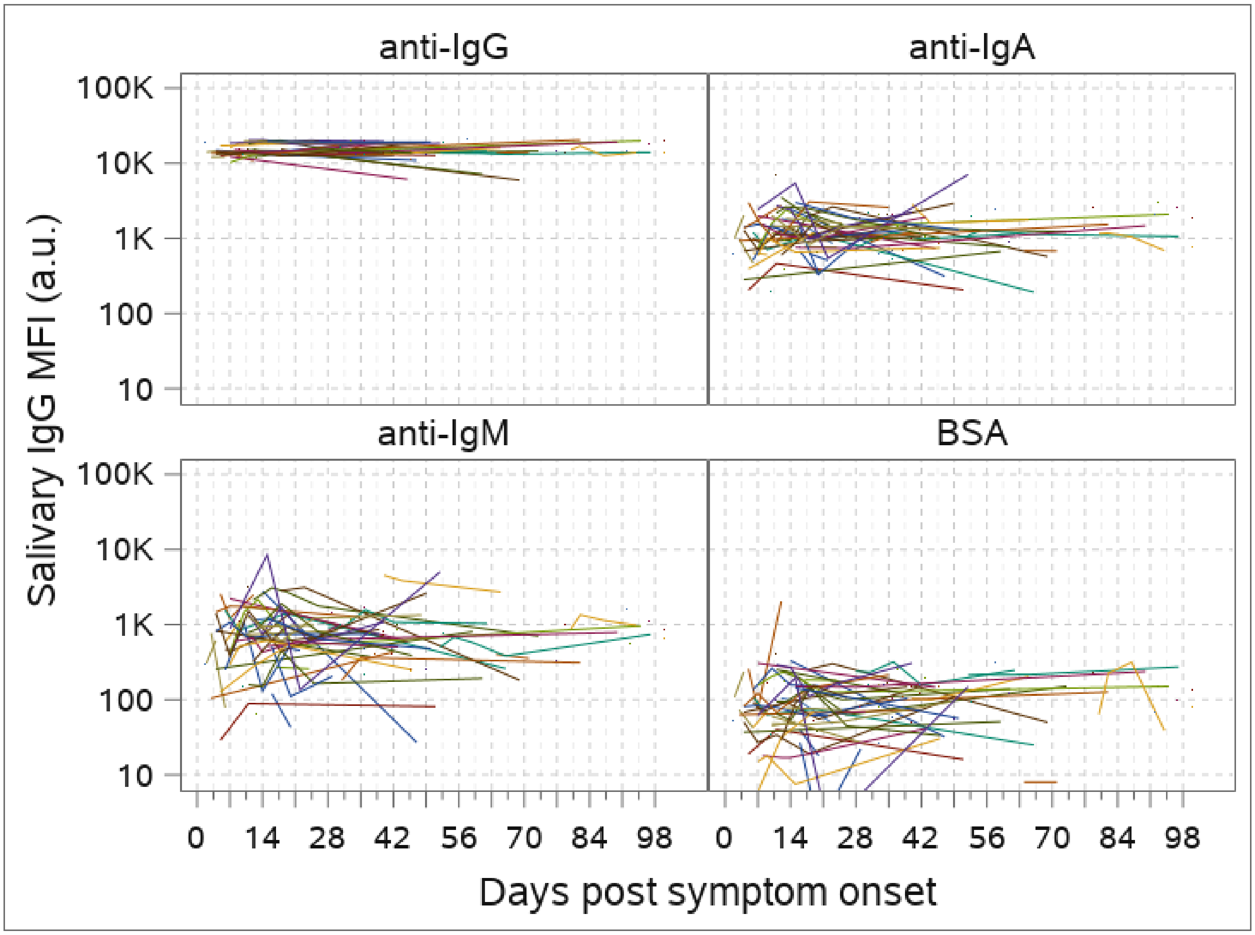


**Supplementary Figure 3. Antibody isotype-specific and bovine serum albumin (BSA)-specific kinetics in oral fluid.** Results shown are from an in-house multiplex SARS-CoV-2 immunoassay based on Luminex technology. Lines connect values from the same participant. To minimize false negatives due to low concentration of total antibody, samples with less than 15 µg/mL total oral fluid IgG are not shown here unless they were also positive for most SARS-CoV-2 specific antigens. MFI, mean fluorescence intensity; a.u. arbitrary units.

**Supplementary Table 1. Days from symptom onset to positive oral fluid SARS-CoV-2-specific IgG.**

| Antigen | Mean | 95% confidence interval |
|---|---|---|
| GenScript N | 10.2 | 9.0 – 11.4 |
| NAC N | 10.0 | 8.8 – 11.2 |
| Sino Biol. RBD (1) | 11.8 | 10.8 – 12.8 |
| Sino Biol. RBD (2) | 9.2 | 7.8 – 10.6 |
| Mt. Sinai RBD | 9.2 | 7.4 – 11.2 |
| Mt. Sinai Spike | 13.6 | 12.4 – 14.8 |
| GenScript RBD | 10.2 | 9.0 – 11.4 |
| Sino Biol. ECD (S1+S2) | 8.0 | 6.6 – 9.4 |

N, nucleocapsid; NAC, Native Antigen Company; RBD, receptor binding domain of spike protein; ECD, spike ectodomain; S1, spike subunit 1; S2, spike subunit 2.

**Supplementary Table 2. Variables selected after multiple imputation and LASSO for inclusion in the Cox proportional hazards model for RT-PCR clearance.**

| Variable | Frequency of selection by Lasso in 20 imputed datasets | Selected for Cox model |
|---|---|---|
| BMI ≥ 25 (referent: BMI < 25) | 20 | Yes |
| COPD or asthma | 20 | Yes |
| Diabetes | 20 | Yes |
| Fever as one of first 3 COVID-19 symptoms | 20 | Yes |
| Immunocompromised | 20 | Yes |
| Female | 19 | Yes |
| Oral fluid Mt Sinai RBD-specific IgG* | 17 | Yes |
| African American or Black race, any ethnicity (referent: White non-Hispanic) | 16 | Yes |
| Oral fluid hCoV NL63 ECD-specific IgG* | 15 | Yes |
| Age > 65 | 14 | No |
| Cancer not in remission | 14 | No |
| Oral fluid hCoV 229E ECD-specific IgG* | 14 | No |
| Oral fluid bovine serum albumin-specific IgG* | 14 | No |
| Ever smoker | 13 | No |
| Plasma anti-RBD ELISA AUC† | 12 | No |
| hCoV HKU1 S1-specific IgG* | 12 | No |
| Plasma anti-Spike ELISA AUC† | 11 | No |
| Oral fluid Mt Sinai Spike-specific IgG* | 11 | No |
| Oral fluid hCoV OC43 HE-specific IgG* | 11 | No |
| Hypertension | 10 | No |
| Autoimmune disease | 9 | No |
| Plasma neutralizing Ab AUC† | 9 | No |
| Oral fluid NAC N-specific IgG* | 9 | No |
| Ct value of 1st RT-PCR (if within 7 days of symptom onset and performed on NeuMoDx) | 8 | No |
| Oral fluid total IgA* | 8 | No |
| Chronic kidney disease | 6 | No |
| Cardiovascular disease (not including hypertension) | 4 | No |
| Oral fluid Sino Biol. ECD (S1 + S2)* | 4 | No |
| Oral fluid GenScript N* | 4 | No |
| Non-White, Non-Black, non-Hispanic (referent: White non-Hispanic) | 3 | Yes |
| Oral fluid total IgM* | 3 | No |
| Hispanic of any race except Black (referent: White non-Hispanic) | 2 | Yes |
| Oral fluid Sino Biol. RBD (2)-specific IgG* | 2 | No |
| Oral fluid Sino Biol. RBD (1)-specific IgG* | 0 | No |
| GenScript RBD (h)-specific IgG* | 0 | No |

*Standardized log mean fluorescence intensity, treated as time-varying variable

[†]Standardized log area under curve (AUC) of antibody titer measured from plasma collected once from each participant at a timepoint 1-3 months after symptom onset, treated as a baseline variable
RBD, spike receptor binding domain; hCoV, human coronavirus; ECD, spike ectodomain; ELISA, enzyme-linked immunosorbent assay; S1, spike subunit 1; HE, hemagglutinin esterase; Ab, antibody; NAC, Native Antigen Company; Ct, cycle threshold; S2, spike subunit 2; N, nucleocapsid.

**Supplementary Table 3. Missing value patterns of longitudinal dataset for Cox Model 1.**

| | Pattern (1 = complete; 0 = missing) | | | | | | | | | | | | | |
|---|---|---|---|---|---|---|---|---|---|---|---|---|---|---|
| **Percent** | (1) | (2) | (3) | (4) | (5) | (6) | (7) | (8) | (9) | (10) | (11) | (12) | (13) | (14) |
| 58% | 1 | 1 | 1 | 1 | 1 | 1 | 1 | 1 | 1 | 1 | 1 | 1 | 1 | 1 |
| 38% | 1 | 1 | 1 | 1 | 1 | 1 | 1 | 0 | 0 | 0 | 0 | 0 | 0 | 0 |
| 1 | 1 | 1 | 1 | 1 | 1 | 1 | 0 | 0 | 0 | 0 | 0 | 0 | 0 | 0 |
| <1 | 1 | 1 | 1 | 1 | 1 | 0 | 1 | 1 | 1 | 1 | 1 | 1 | 1 | 1 |
| <1 | 1 | 1 | 1 | 1 | 0 | 1 | 1 | 1 | 1 | 1 | 1 | 1 | 1 | 1 |
| <1 | 1 | 1 | 1 | 1 | 1 | 0 | 1 | 0 | 0 | 0 | 0 | 0 | 0 | 0 |
| <1 | 0 | 0 | 0 | 0 | 0 | 0 | 0 | 0 | 0 | 0 | 0 | 0 | 0 | 0 |
| <1 | 0 | 0 | 0 | 0 | 1 | 0 | 0 | 0 | 0 | 0 | 0 | 0 | 0 | 0 |
| <1 | 1 | 1 | 1 | 1 | 0 | 0 | 1 | 0 | 0 | 0 | 0 | 0 | 0 | 0 |
| <1 | 1 | 1 | 1 | 1 | 0 | 1 | 0 | 0 | 0 | 0 | 0 | 0 | 0 | 0 |
| <1 | 1 | 1 | 1 | 1 | 0 | 1 | 0 | 1 | 1 | 1 | 1 | 1 | 1 | 1 |
| <1 | 1 | 1 | 1 | 1 | 0 | 1 | 1 | 0 | 0 | 0 | 0 | 0 | 0 | 0 |
| <1 | 1 | 1 | 1 | 1 | 1 | 1 | 1 | 1 | 1 | 1 | 1 | 1 | 1 | 0 |
| 100% | | | | | | | | | | | | | | |

**Variables are (1) copd; (2) cvd; (3) diabetes; (4) hypertension; (5) smoking; (6) BMI group; (7) fever; (8) std_ln(_229e); (9) std_ln(gen_N); (10) std_ln(hku1); (11) std_ln(lx_n); (12) std_ln(nl63); (13) std_ln(oc43); (14) std_ln(sino_rbd)**

58% of data were complete. 38% of data were missing on variables (7)-(14) above.

**Supplementary Table 4. Logistic regression of missingness of Table 2 variables 7-14 shows missingness did not depend on any known variables and the data appeared most likely missing at random (MAR).**

**Supplementary Table 3. Logistic regression of missingness**

| Variables | Odds ratio | Std. Err. | Z | P>\|Z\| | 95% CI. lower | 95% CI. upper |
|---|---|---|---|---|---|---|
| Age | 1.08 | 0.26 | 0.33 | 0.74 | 0.68 | 1.72 |
| Sex | 1.18 | 0.24 | 0.82 | 0.41 | 0.80 | 1.75 |
| COPD | 0.80 | 0.23 | -0.78 | 0.44 | 0.45 | 1.42 |
| CVD | 1.60 | 0.43 | 1.74 | 0.08 | 0.94 | 2.70 |
| Diabetes | 1.17 | 0.32 | 0.57 | 0.57 | 0.68 | 2.02 |
| Hypertension | 1.21 | 0.25 | 0.94 | 0.35 | 0.81 | 1.81 |
| Smoking | 0.91 | 0.21 | -0.40 | 0.69 | 0.58 | 1.44 |
| BMI group | 1.30 | 0.33 | 1.03 | 0.30 | 0.79 | 2.14 |
| Fever | 0.75 | 0.16 | -1.36 | 0.18 | 0.49 | 1.14 |

**Supplementary Table 5. Sensitivity analysis: Cox proportional hazards model for RT-PCR clearance excluding participants who cleared ≥ 75 days after symptom onset.**

| Variable | HR | 95% CI | p value |
|---|---|---|---|
| BMI ≥ 25 $kg/m^2$ (referent: BMI < 25) | 0.32 | 0.15 – 0.70 | 0.004 |
| COPD or asthma | 3.58 | 1.43 – 8.91 | 0.006 |
| Diabetes | 1.99 | 0.85 – 4.69 | 0.113 |
| Immunocompromised | 0.46 | 0.15 – 1.36 | 0.159 |
| Fever as one of first 3 COVID-19 symptoms | 1.41 | 0.71 – 2.78 | 0.325 |
| Female | 0.72 | 0.38 – 1.36 | 0.317 |
| Mt Sinai S-specific oral fluid IgG* | 0.64 | 0.40 – 1.02 | 0.060 |
| hCoV OC43 HE-specific oral fluid IgG* | 0.83 | 0.50 – 1.38 | 0.470 |
| hCoV 229E ECD-specific oral fluid IgG* | 1.26 | 0.83 – 1.92 | 0.272 |

*Standardized log mean fluorescence intensity, treated as a time-varying variable.
HR, hazard ratio; CI, confidence interval, S, SARS-CoV-2 spike protein; hCoV, human coronavirus; HE, hemagglutinin esterase; ECD, ectodomain

**Supplementary Methods**

*Oral fluid immunoassay antigens and total IgG measurement.* The immunoassay described in the main methods section included SARS-CoV-2 nucleocapsid (N), receptor binding domain (RBD), spike (S) antigens, SARS, MERS, RSV, human coronavirus E229, NL63, HKU1, and OC43 antigens in addition to control antibodies and proteins (BSA, anti-human IgG, IgM, IgA antibody). The total IgG concentration in oral fluid was determined using Salimetrics Salivary Human Total IgG ELISA Kits according to the manufacturer's instructions. Cutoffs for each SARS-CoV-2 antigen were defined as the average net (blank-subtracted) median fluorescence intensity of pre-COVID-19 era oral fluid samples plus three standard deviations (SD).

*Oral fluid IgG kinetics.* To minimize false negatives, datapoints used in Suppl. Fig. 1 and Suppl. Table 2 are those with a minimum of 15 µg/mL total IgG by ELISA or those with a sum of signal to cutoff to seven SARS-CoV-2 N, RBD, and S antigens greater than 6. 72 of 292 total data points were excluded using this criteria, with the majority in the first 7 days post-symptom onset. The mean number of days and 95% confidence intervals from symptom onset until the detection of oral fluid IgG to each SARS-CoV-2-specific antigen were calculated using a linear regression of data points ≤ 20 days post symptom onset and determining mean number of days and 95% CI when the regression intercepted the cutoff using SAS software.

*Cox models.* Associations between variables related to the host, illness, or immunity and clearance of upper respiratory tract SARS-CoV-2 viral RNA as measured by RT-PCR was determined via Cox proportional hazards models[3] after multiple imputation and model selection by least absolute shrinkage and selection operator (LASSO). A complete list of covariates and their status as time-independent or time varying for Cox model 1 may be found in Supplementary Table 2. Time to RT-PCR clearance was defined as number of days from symptom onset to the midpoint between the last positive RT-PCR test and the subsequent negative RT-PCR test. Patterns of missing data were investigated by generating a missing value patterns table (Suppl. Table 3) and parallel blox plots characterizing missing values using the visualization and imputation of missing values (VIM) package[4] in R[5]. Logistic regression was performed on the most frequent missing patterns to confirm that the data were missing at random (Suppl. Table 4). 20 datasets were imputed using predictive mean matching for univariate imputation and chained equation (MICE) for multivariate imputation using the 'mice' R package[6]. Model selection was performed using least absolute shrinkage and selection operator (LASSO) on each of the 20 imputed datasets using the 'glmnet' R package[7]. 9 variables were included in the model that were selected no less than 15 times out of the 20 model selections (Supp. Table 2). Using the pooled imputed dataset, we used Cox model 1 to identify what covariates were associated with RT-PCR clearance and estimated the hazard ratios of clearance and 95% confidence intervals of each (Fig. 5). $Cox\ Model\ 1: h\big(t; X, Z(t)\big) = h_0(t) exp\{aX + bZ(t)\}$, where h denotes hazard, $h_0$ denotes baseline hazard, X denotes time-independent covariates, Z denotes time-dependent covariates, and t denotes time.

Sensitivity analysis using Cox regression model 1 was performed with the method described above but excluding participants who had length of positive RT-PCR ≥ 75 days (Suppl. Table 5).

To determine whether the time from symptom onset to first detection of salivary SARS-CoV-2-specific IgG is associated with RT-PCR clearance, we performed model selection using LASSO in a second Cox model (model 2). Hazard ratios of clearance and 95% confidence intervals were estimated. $Cox\ Model\ 2: h\big(t; X, Z(t)\big) = h_0(t)\exp(aX + (b + cS)Z(t))$ where h denotes hazard, $h_0$ denotes baseline hazard, X denotes time-independent covariates, S denotes time to first detection of oral fluid SARS-CoV-2-specific IgG above cutoff, Z(t) = I(S<=t).
